# Perceived stress and associated societal, community, interpersonal, and individual factors in adults with post-COVID-19 condition: the PRIME study

**DOI:** 10.1101/2025.11.24.25340916

**Authors:** Senne MCE Wijnen, Céline JA van Bilsen, Demi ME Pagen, Annemarie Koster, Christian JPA Hoebe, Nicole HTM Dukers-Muijrers

## Abstract

**Background:** Post-COVID-19 condition (PCC) is a global health concern with a profound impact on physical, mental, and social health, potentially leading to stress. This study compared perceived stress levels between adults with PCC and those without PCC (non-PCC).

Additionally, environmental (societal, community, interpersonal) and individual factors were explored for their association with stress.

**Methods:** Cross-sectional questionnaire data from participants with SARS-CoV-2 infection were obtained from the prospective PRIME post-COVID cohort (2022). PCC was defined as feeling unrecovered (≥ 3 months) after SARS-CoV-2 infection. Associations between societal, community, interpersonal and individual factors and the outcome perceived stress (measured by the Perceived Stress Scale 14; PSS-14) were assessed in PCC and non-PCC using logistic regressions, adjusted for gender and age.

**Results:** In total, 3275 participants were included (PCC: n=1044, non-PCC: n=2231). PCC had higher perceived stress scores (mean± SE: 20.75± 0.27) than non-PCC (mean± SE: 15.65± 0.19), adjusted for age and gender (p<0.001). Factors associated with more perceived stress included lower neighborhood livability and cohesion (societal level), higher absenteeism and presenteeism (community level), less social support, less dense and diverse social network (interpersonal level), being female, having co-morbidities, post-exertional malaise, orthostatic intolerance, loneliness, and lower coping score (individual level). These factors and their effect-sizes were largely comparable between PCC and non-PCC, but nearly all were more prevalent in PCC compared to non-PCC group.

**Conclusion:** Adults with PCC perceived higher stress levels compared to adults without PCC, which may be attributable to higher prevalences of a multitude of stress-associated environmental and individual factors.

## 1.0 Introduction

Since the emergence of SARS-CoV-2 in December 2019, the World Health Organization has reported over 775 million cases of acute SARS-CoV-2 infections globally by June 2024 [1]. After an acute SARS-CoV-2 infection, people may have various persistent or intermitting, often debilitating, symptoms named post-COVID-19 condition (PCC) or long COVID. The National Academies Sciences Engineering and Medicine (NASEM) defines PCC as an infection-associated chronic condition that occurs after SARS-CoV-2 infection and is present for at least 3 months as a continuous, relapsing and remitting, or progressive disease state that affects one or more organ systems [2]. Globally, at least an estimated 65 million people were affected by PCC [3], and in the Netherlands at least 1 in 6 adults was estimated to have developed PCC [4]. Hence, PCC constitutes a global health concern of considerable magnitude.

PCC has a profound multifaceted impact on an individual. Studies have demonstrated that people with PCC are more impaired in their daily activities and report lower quality of life and a worse perceived health [5, 6]. On the social and medical domain, PCC may be accompanied by difficulties in maintaining or building new social relationships [7, 8], and finding adequate support from healthcare providers [9]. Furthermore, having PCC may have major occupational implications, as previous studies showed that about 20% of people with PCC were unable to work due to their illness [10, 11]. PCC has a clear single biological cause that can lead to various pathophysiological mechanisms capable of harming nearly any organ or tissue [12–14]. This results in multifaceted consequences encompassing the individual physical, mental, and social health, interpersonal relationships, community dynamics, and the broader society, following the socio-ecological model.

Living with PCC and its consequences may cause (di)stress, meaning that the impact of PCC (and its consequences) may exceed the individual’s capacity to effectively cope with the situation. This notion is supported by studies showing higher stress levels in PCC [15–17].

Having a chronic condition may lead to distress as demonstrated for example in diabetes patients [18]. Furthermore, stress is a known risk factor for a wide variety of health conditions, such as Alzheimer’s disease, rheumatoid arthritis, obesity, and cardiovascular diseases [19–22]. This highlights the importance of science, clinical practice, and society understanding stress in PCC.

Factors that are generally associated with stress, such as living in a deprived neighborhood, unemployment, younger age, being women, lower educational level, fatigue, anxiety, depression, less coping, and smoking, have been extensively studied across different populations [23–25]. However, studies identifying factors associated with perceived stress in adults with PCC are lacking. In the current observational study, we compared perceived stress between adults with and without PCC, and further compared stress levels across more detailed subgroups: persistent, former, new-onset PCC and those who never had PCC. In addition, potential associations were assessed between the outcome perceived stress and a wide variety of factors (at societal, community, interpersonal, and individual level) in adults with current PCC. This analysis was also conducted in adults without PCC, with the aim to identify possible stress-associated factors specific to PCC, with the goal of informing policy and care.

## 2.0 Methods

### 2.1 Ethics statement

The Medical Ethical Committee of Maastricht University Medical Center+, Maastricht Netherlands waived this study (METC2021-2884), as the Medical Research Involving Human Subjects Act (WMO) did not apply to this study. The study was conducted in accordance with the principles of the Declaration of Helsinki, and is registered at <u>ClinicalTrials.gov</u> Protocol Registration and Results System (<u>NCT05128695</u>). Participants gave written electronic informed consent.

### 2.2 Study design and participants

The Prevalence, Risk factors, and Impact Evaluation (PRIME) post-COVID study is an observational cohort study [26]. Adult women and men (18 years and older), who tested for SARS-CoV-2 at the local public health service (GGD) (South Limburg, The Netherlands) between 1 June 2020 and 1 November 2021, were invited for the baseline questionnaire (17 November 2021- 9 January 2022). 12,453 individuals completed the baseline questionnaire and were invited to complete the follow-up questionnaire (21 July 2022 - 11 September 2022), in which perceived stress was measured. Participants, who completed the follow-up questionnaire (all relevant questions), were the intended invitee with sufficient certainty (match on gender, age and test result), and previously tested positive for SARS-CoV-2 (first positive test more than 3 months prior to follow-up, based on the national public health medical registry and self-report), were included in the current analysis. Data were self-reported using digital questionnaires and obtained by linking to census-tract data, whereas SARS-CoV-2 test results were obtained from the public health medical registry. The MWM2 application of market research platform Crowdtech (ISO 27001 certified) was used for data collection.

### 2.3 Post-COVID-19 condition (PCC)

Participants were defined as having PCC when they felt unrecovered after their SARS-CoV-2 infection. This definition of PCC was chosen since this reflects the population who would seek (healthcare) support and is in line with previous studies [4, 27–31]. PCC status at baseline (7-9 months prior to follow-up) was taken to define persistent PCC (PCC both at baseline and follow-up), new onset PCC (PCC only at follow-up), former PCC (PCC at baseline and recovered in follow-up), and never PCC (no PCC at baseline and follow-up) (Fig 1).

**Fig. 1.**
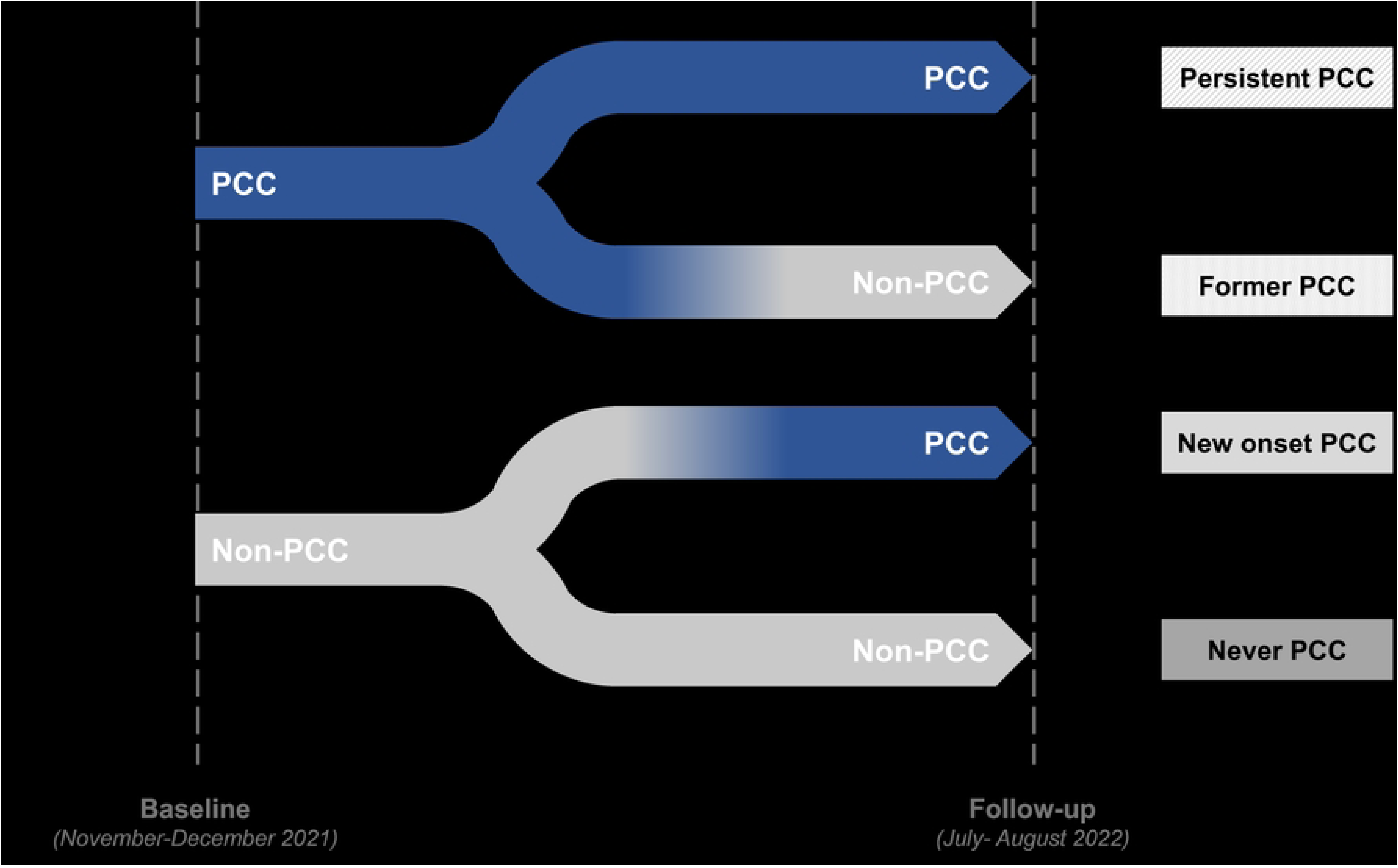
Classification of persistent, former, new onset and never post-COVID-19 condition (PCC) based on baseline and follow-up PCC status

### 2.4 Perceived stress

The main outcome of this study was perceived stress in the past month, measured by the Dutch version of the validated perceived stress scale 14 (PSS-14) [32]. The scale consisted of 14 items, scored on a 5-point Likert scale ranging from 0 (“never”) to 4 (“very often”) (S1 table). The total PSS-14 score was obtained by inverting the positively stated items, and subsequently summing across the 14 items. The resulting score ranged from 0-56, with higher scores indicating higher perceived stress. Cronbach alpha showed good internal consistency (`α`=0.876) within the current sample.

As PSS-14 is not a diagnostic tool, there were no predetermined cut-off points. Hence, we used the median score of the total current sample as cut-off for logistic regression analysis. The use of a median cut-off is in line with previous studies [33–36].

### 2.5 Socio-ecological model

The current study aimed to explore a wide range of factors from four levels, each of which has previously been shown to contain factors associated with perceived stress [23–25]. These levels were based on a modified version of Bronfenbrenner’s socio-ecological model [37–39] (Fig 2). An extended description of some variables is included (S2 table).

**Fig. 2.**
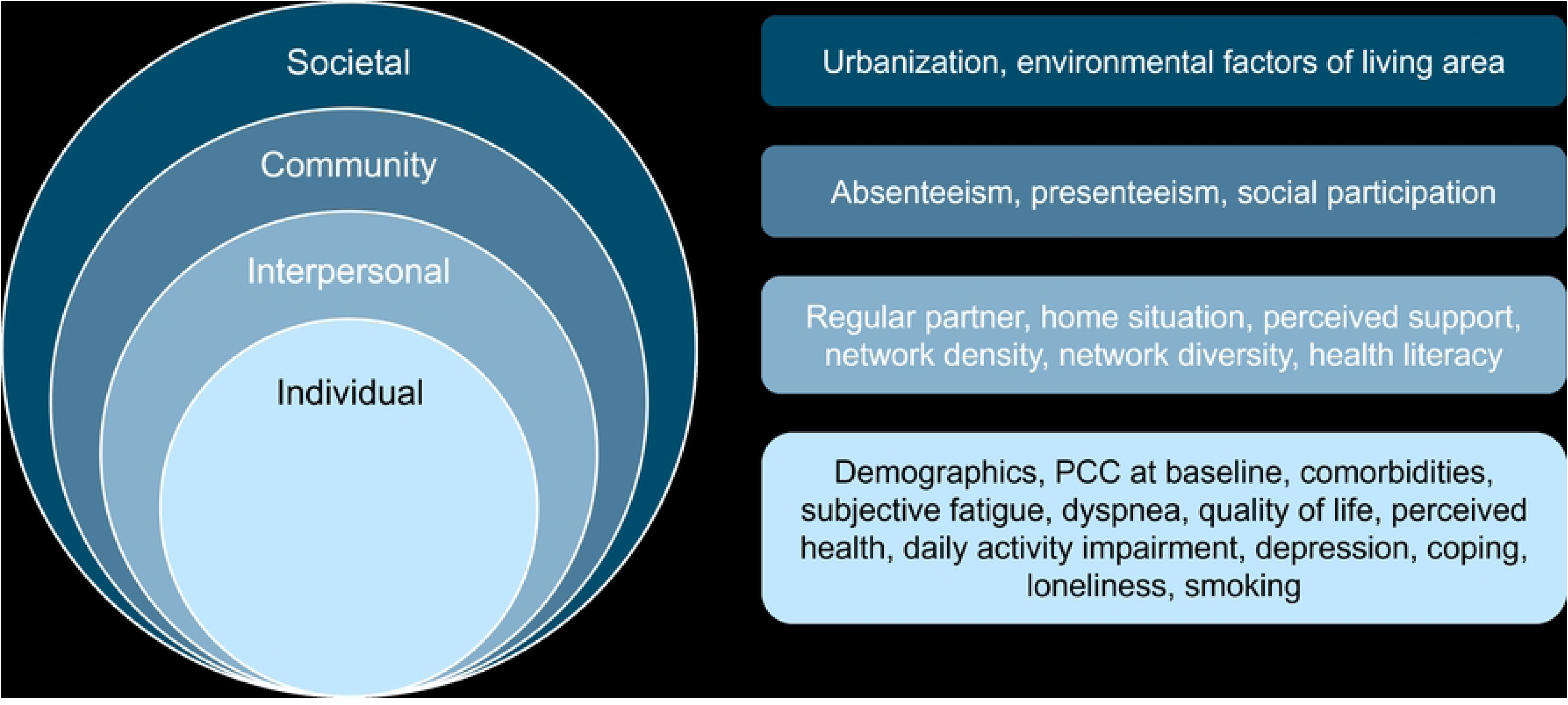
Overview of factors studied for their potential association with perceived stress using the socio-ecological model

### 2.6 Societal factors (census tract data)

Urbanization of living area was determined by linking the participants’ 4-digit postal code to urbanization data of Statistics Netherlands (CBS). The remaining factors of living area were obtained from Geoscience and Health Cohort Consortium (GECCO) [40, 41], and were linked to individual participants based on their 4- and 6-digit postal codes. These factors included: green space density, daily noise, air pollution, financial well-being, social cohesion (indirect measure) and livability, which indicates the extent to which living environment is line with the needs and conditions of residents (S2 table). Some participants could not be linked to living area data, because of postal codes less than five inhabitants or missing postal code. Consequently, they were excluded from analyses of environmental factors of living area, but included in those of other factors.

### 2.7 Community factors (self-reported data)

Absenteeism and presenteeism were determined using the Work Productivity and Activity Impairment - General Health (WPAI-GH) questionnaire [42]. Absenteeism was defined as the percentage of work time missed due to health problems in the past 7 days. Presenteeism represented the percentage of impairment while working due to health problems in the past 7 days, with higher percentages indicating less productivity. Unemployed participants were categorized separately. Furthermore, social participation was evaluated by reporting their membership of sport associations or walking group, cultural organizations (music, theater, dance or traditional regional association) and/or internet group.

### 2.8 Interpersonal factors (self-reported data)

Perceived social support from family, friends and significant others was measured using the Multidimensional Scale of Perceived Social Support (MSPSS) (S2 table). Four domains of the Health Literacy Questionnaire (HLQ) were utilized to measure whether participants had sufficient information to manage health (HLQ2), received social support for health (HLQ4), were able to actively engage with healthcare providers (HLQ6) and were able to navigate the healthcare system (HLQ7) (S2 table). Network density was measured by asking whether their best friend(s) know their family (yes/no). The diversity of the participants’ network was assessed by questioning “Who are the people important to you or from whom you receive support/ to whom you provide support?”, allowing to distinguish between family, friends and acquaintances. Additional variables at interpersonal level involved having a steady partner (yes/no) and who the participants live with (alone/child(ren)/other residents).

### 2.9 Individual factors (self-reported data)

#### 2.9.1 Demographics and lifestyle

Gender (men/women), age (18-40/41-60/ >60 years), educational level (low/medium/high), and smoking status (never/former/current) were obtained.

#### 2.9.2 Physical health

Comorbidities such as diabetes, cardiovascular diseases, asthma, COPD, fibromyalgia (yes/no) were obtained, as well as the number of conditions (0-29) (categorized into 0, 1-2 and >2). Post-exertional malaise (PEM) was assessed using the 5-item DePaul Symptom Questionnaire (DSQ-PEM), whereas orthostatic intolerance (OI) was measured with four items selected from the DSQ-2 according to the chronic fatigue syndrome (ME/CFS) classifications. Subjective fatigue and dyspnea were determined by the subscale of Checklist Individual Strength (CIS) and by the modified Medical Research Council (mMRC) dyspnea scale, respectively. Quality of life (QoL) was reported using the EuroQol five-dimensions with five levels (EQ5D-5L), which contained five health domains (mobility, self-care, daily activities, pain/discomfort and anxiety/depression) (S2 table). The EQ5D-5L included the vertical visual analogue scale (EQVAS) to measure perceived health (range 0-100). As part of the WPAI, participants were asked to rate to what extent their health problems affected their ability to do their regular daily activities (except work) in the past 7 days, from 0 (no effect) to 10 (major effect). Daily activity impairment was expressed as a percentage with higher percentages indicating a greater impairment [42].

#### 2.9.3 Mental and social health

Depression was classified according to the Patient Health Questionnaire 9 (PHQ-9). Moreover, coping was determined using ten items from the original 36-items Cognitive Emotion Regulation Questionnaire (CERQ), and two previously self-generated items.

Loneliness was measured by the 6-item De Jong Gierveld Loneliness scale (S2 table).

### 2.10 Statistical analysis

Differences in participant characteristics between adults with PCC and without PCC (non-PCC) were analyzed using descriptives and logistic regression (outcome variable: PCC versus non-PCC, independent variables: participant characteristics) adjusted for age and gender.

The mean PSS-14 stress scores were compared between current PCC and non-PCC, as well as between never, former, new onset and persistent PCC by performing an ANCOVA adjusted for age and gender.

Each factor from the socio-ecological model was separately evaluated for its association with the outcome perceived stress (above the median versus ≤ median) using logistic regression analyses adjusted for age and gender. Analyses were conducted separately for the PCC and non-PCC groups. Subsequently, the data of the two groups were combined and possible heterogeneity of effects was tested, by including interaction terms between PCC group (PCC or non-PCC) and socio-ecological factors.

Prior to multivariable analysis, multicollinearity tests were performed using the Variation Inflation Factor (VIF) and Spearman correlations, with VIF> 5, ρ> 0.800 or ρ< -0.800 leading to exclusion. We hypothesized that stress may differ between PCC and non-PCC. In a final analysis, we evaluated whether the socio-ecological levels contributed to a difference in stress (if any). Therefore, we conducted multivariable logistic regressions with perceived stress as outcome measure, adjusted for age and gender. Starting with a basic model that included only PCC group as independent variable, each subsequent model incorporated an additional socio-ecological level, beginning with societal and ending with individual level. Only factors that were found significantly associated with perceived stress were included in these models. The percentage change in the odds ratio of perceived stress for PCC groups, with the addition of each socio-ecological level, suggested which levels might explain a possible difference in perceived stress between PCC and non-PCC. Analyses were performed using Statistical Package for Social Sciences (SPSS; version 27.0, IBM, Armonk, USA). A p-value of <0.05 was considered statistically significant.

## 3.0 Results

The study population consisted of 3275 individuals, of whom 1044 had PCC (779 persistent, 265 new onset), and 2231 were without PCC (388 former PCC, 1843 never had PCC) (Fig 3). In the PCC group, 60.8% were women, 48.9% were 41-60 years old and 40.5% were older than 60 years, compared to 57.0%, 42.1% and 42.5% in the non-PCC group, respectively.

**Fig. 3.**
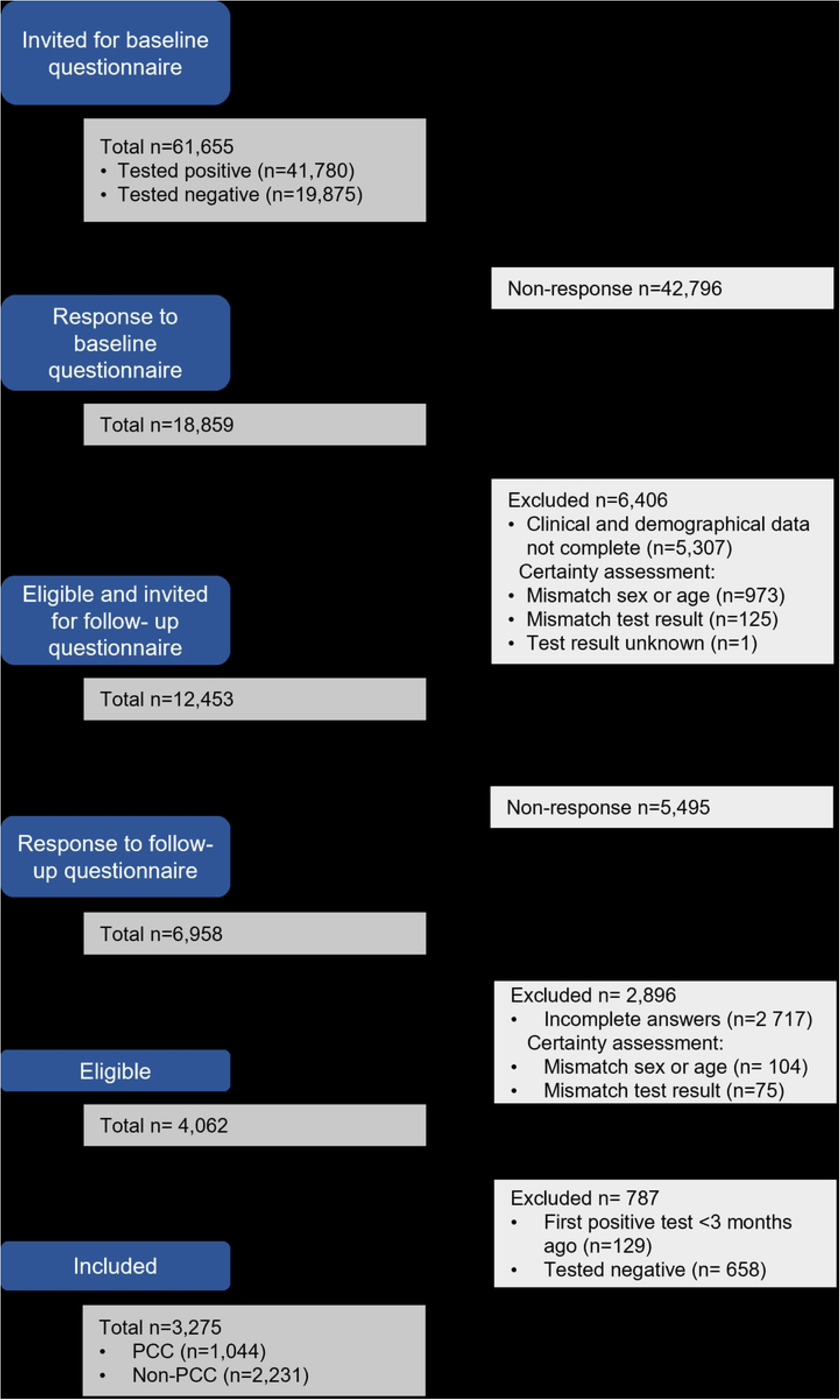
Flowchart of participants from the PRIME study included in the current analysis

### 3.1 Comparison of characteristics between PCC and non-PCC

At the societal level, adults with PCC showed lower livability scores, and lived in neighborhoods with less cohesion and a worse financial well-being, compared to the non-PCC group (Table 1). At the community level, participants with PCC had more absenteeism and presenteeism due to health problems, and were less often member of sport association or walking group. At the interpersonal level, adults with PCC perceived less social support from family, friends and significant others, had a less dense and less diverse social network, and lower health literacy.

**Table 1.** Characteristics of the participants from the PRIME study with post-COVID-19 condition (PCC) and without post-COVID-19 condition (non-PCC).

| | **PCC**<br>n= 1044<sup>a</sup><br>n (%) or<br>mean ± SD | **Non-PCC**<br>n= 2231<sup>a</sup><br>n (%) or<br>mean ± SD | **p-value**<br>(adjusted for age<br>and gender) |
| --- | --- | --- | --- |
| **Societal level** |  |  |  |
| **Urbanization of living area** |  |  | **0.153** |
| Not urban | 182 (17.4%) | 390 (17.5%) |  |
| Little urban | 238 (22.8%) | 548 (24.6%) |  |
| Moderately urban | 255 (24.4%) | 465 (20.8%) |  |
| Strongly urban | 369 (35.3%) | 828 (37.1%) |  |
| **Green space density, radius 500m of living area (%)** | 56.8 ± 12.1<br>(n=1026) | 57.5 ± 12.1<br>(n=2203) | **0.107** |
| **Daily noise of living area (dB)** | 52.2 ± 5.3<br>(n=1029) | 51.9 ± 5.2<br>(n=2205) | **0.088** |
| **Air pollution PM10 of living area (per m3)** | 15.1 ± 0.4<br>(n=1029) | 15.1 ± 0.4<br>(n=2205) | **0.448** |
| **Air pollution PM2.5 of living area (per m3)** | 8.2 ± 0.2<br>(n=1029) | 8.2 ± 0.2<br>(n=2205) | **0.380** |
| **Air pollution NO2 of living area (per m3)** | 12.2 ± 1.9<br>(n=1029) | 12.2 ± 2.0<br>(n=2205) | **0.925** |
| **Financial well-being, average percentile of living area** | 47.4 ± 9.0<br>(n=1027) | 48.2 ± 9.3<br>(n=2204) | **0.025** |
| **Livability score of living area** | 4.09 ± 0.11<br>(n=993) | 4.12 ± 0.12<br>(n=2119) | **<0.001** |
| **Social cohesion of living area** | 0.010 ± 0.039<br>(n=993) | 0.014 ± 0.041<br>(n=2119) | **0.028** |
| **Community level** |  |  |  |
| **Absenteeism due to health issues** |  |  | **<0.001** |
| Employed, absenteeism 0% | 449 (43.0%) | 1222 (54.8%) |  |
| Employed, absenteeism >0% ≤33% | 27 (2.6%) | 31 (1.4%) | |
| Employed, absenteeism >33% ≤66% | 50 (4.8%) | 32 (1.4%) | |
| Employed, absenteeism >66% | 89 (8.5%) | 40 (1.8%) |  |
| Employed, did not work past week for reasons other than health issues | 53 (5.1%) | 137 (6.1%) |  |
| Not employed | 376 (36.0%) | 769 (34.5%) |  |
| **Presenteeism due to health issues** |  |  | **<0.001** |
| Employed, presenteeism 0% | 95 (9.1%) | 834 (37.4%) |  |
| Employed, presenteeism >0% ≤33% | 144 (13.8%) | 284 (12.7%) | |
| Employed, presenteeism >33% ≤66% | 163 (15.6%) | 110 (4.9%) | |
| Employed, presenteeism >66% | 157 (15.0%) | 65 (2.9%) |  |
| Employed, did not work past week because of health issues | 56 (5.4%) | 32 (1.4%) |  |
| Employed, did not work past week for reasons other than health issues | 53 (5.1%) | 137 (6.1%) |  |
| Not employed | 376 (36.0%) | 769 (34.5%) |  |
| **Member of sport association or walking group** |  |  | **<0.001** |
| No | 826 (79.1%) | 1550 (69.5%) |  |
| Yes | 218 (20.9%) | 681 (30.5%) |  |
| **Member of music, theater, dance or traditional regional association** |  |  | **0.189** |
| No | 934 (89.5%) | 1954 (87.6%) |  |
| Yes | 110 (10.5%) | 277 (12.4%) |  |
| **Member of a group at internet (e.g. Facebook)** |  |  | **0.139** |
| No | 986 (94.4%) | 2135 (95.7%) |  |
| Yes | 58 (5.6%) | 96 (4.3%) |  |
| **Interpersonal level** |  |  |  |
| **Regular/steady partner** |  |  | 0.576 |
| No | 167 (16.0%) | 387 (17.3%) |  |
| Yes | 877 (84.0%) | 1844 (82.7%) |  |
| **Home situation** |  |  | 0.725 |
| Living with other residents, without children | 547 (52.4%) | 1201 (53.8%) |  |
| Living with children and other residents | 267 (25.6%) | 525 (23.5%) |  |
| Living with children only | 53 (5.1%) | 125 (5.6%) |  |
| Living alone | 177 (17.0%) | 380 (17.0%) |  |
| **Social support from family** | 22.5 ± 6.5 | 23.7 ± 5.6 | <0.001 |
| **Social support from friends** | 22.3 ± 6.6 | 23.6 ± 5.6 | <0.001 |
| **Social support from significant others** | 24.2 ± 6.0 | 24.6 ± 5.6 | 0.030 |
| **Social support for health** | 3.08 ± 0.62 | 3.27 ± 0.54 | <0.001 |
| **Network density** |  |  | 0.012 |
| Friends do not know my family | 275 (26.3%) | 499 (22.4%) |  |
| Friends do know my family | 769 (73.7%) | 1732 (77.6%) |  |
| **Network diversity** |  |  | 0.002 |
| Family, friends and acquaintances | 463 (44.3%) | 1126 (50.5%) |  |
| Only family and friends, no acquaintances | 357 (34.2%) | 721 (32.2%) |  |
| Only family and acquaintances, no friends | 47 (4.5%) | 102 (4.6%) |  |
| No family, only friends and acquaintances | 66 (6.3%) | 111 (5.0%) |  |
| Only family, no friends or acquaintances | 111 (10.6%) | 171 (7.7%) |  |
| **Having sufficient information to manage health** | 2.78 ± 0.57 | 3.27 ± 0.54 | <0.001 |
| **Ability to actively engage with healthcare providers** | 3.93 ± 0.88 | 4.52 ± 0.62 | <0.001 |
| **Navigating the healthcare system** | 3.58 ± 0.90 | 4.24 ± 0.70 | <0.001 |
| **Individual level** |  |  |  |
| **Demographics** |  |  |  |
| **Gender** |  |  | 0.028 |
| Men | 409 (39.2%) | 959 (43.0%) |  |
| Women | 635 (60.8%) | 1272 (57.0%) |  |
| **Age** |  |  | <0.001 |
| 18-40 years | 111 (10.6%) | 343 (15.4%) |  |
| 41-60 years | 510 (48.9%) | 939 (42.1%) |  |
| > 60 years | 423 (40.5%) | 949 (42.5%) |  |
| **Educational level** |  |  | <0.001 |
| Low educational level | 254 (24.3%) | 371 (16.6%) |  |
| Medium educational level | 393 (37.6%) | 740 (33.2%) |  |
| High educational level | 397 (38.0%) | 1120 (50.2%) |  |
| **Physical health** |  |  |  |
| **PCC at baseline (7-9 months ago)** |  |  | <0.001 |
| No | 265 (25.4%) | 1843 (82.6%) |  |
| Yes | 779 (74.6%) | 388 (17.4%) |  |
| **Comorbidities** |  |  | <0.001 |
| No comorbidities | 269 (25.8%) | 1092 (48.9%) |  |
| 1-2 comorbidities | 517 (49.5%) | 1000 (44.8%) |  |
| >2 comorbidities | 258 (24.7%) | 139 (6.2%) |  |
| **Diabetes** |  |  | <0.001 |
| No | 950 (91.0%) | 2124 (95.2%) |  |
| Yes | 94 (9.0%) | 107 (4.8%) |  |
| **Cardiovascular diseases (heart diseases, stroke, thrombosis, peripheral vascular disease)** |  |  | <0.001 |
| No | 910 (87.2%) | 2086 (93.5%) |  |
| Yes | 134 (12.8%) | 145 (6.5%) |  |
| **Asthma** |  |  | **<0.001** |
| No | 929 (89.0%) | 2141 (96.0%) |  |
| Yes | 115 (11.0%) | 90 (4.0%) |  |
| **COPD** |  |  | **<0.001** |
| No | 979 (93.8%) | 2181 (97.8%) |  |
| Yes | 65 (6.2%) | 50 (2.2%) |  |
| **Fibromyalgia** |  |  | **<0.001** |
| No | 962 (92.1%) | 2195 (98.4%) |  |
| Yes | 82 (7.9%) | 36 (1.6%) |  |
| **PEM** |  |  | **<0.001** |
| No | 529 (50.7%) | 2003 (89.8%) |  |
| Yes | 515 (49.3%) | 228 (10.2%) |  |
| **OI** |  |  | **<0.001** |
| No | 726 (69.5%) | 2081 (93.3%) |  |
| Yes | 318 (30.5%) | 150 (6.7%) |  |
| **Subjective fatigue** |  |  | **<0.001** |
| Normal fatigue | 175 (16.8%) | 1548 (69.4%) |  |
| Mild fatigue | 259 (24.8%) | 419 (18.8%) |  |
| Severe fatigue | 610 (58.4%) | 264 (11.8%) |  |
| **Dyspnea** |  |  | **<0.001** |
| Grade 0 | 472 (45.2%) | 1848 (82.8%) |  |
| Grade 1 | 386 (37.0%) | 334 (15.0%) |  |
| Grade 2 | 149 (14.3%) | 42 (1.9%) |  |
| Grade 3 | 26 (2.5%) | 6 (0.3%) |  |
| Grade 4 | 11 (1.1%) | 1 (0.0%) |  |
| **EQ5D – domain mobility** |  |  | **<0.001** |
| No problems | 496 (47.5%) | 1779 (79.7%) |  |
| Mild problems | 487 (46.6%) | 412 (18.5%) |  |
| Severe problems | 61 (5.8%) | 40 (1.8%) |  |
| **EQ5D – domain self-care** |  |  | **<0.001** |
| No problems | 901 (86.3%) | 2168 (97.2%) |  |
| Mild problems | 138 (13.2%) | 59 (2.6%) |  |
| Severe problems | 5 (0.5%) | 4 (0.2%) |  |
| **EQ5D – domain daily activities** |  |  | **<0.001** |
| No problems | 356 (34.1%) | 1846 (82.7%) |  |
| Mild problems | 594 (56.9%) | 361 (16.2%) |  |
| Severe problems | 94 (9.0%) | 24 (1.1%) |  |
| **EQ5D – domain pain/discomfort** |  |  | **<0.001** |
| No problems | 199 (19.1%) | 1211 (54.3%) |  |
| Mild problems | 744 (71.3%) | 990 (44.4%) |  |
| Severe problems | 101 (9.7%) | 30 (1.3%) |  |
| **EQ5D – domain anxiety/depression** |  |  | **<0.001** |
| No problems | 596 (57.1%) | 1846 (82.7%) |  |
| Mild problems | 416 (39.8%) | 370 (16.6%) |  |
| Severe problems | 32 (3.1%) | 15 (0.7%) |  |
| **EQ5D index/ QoL** | 0.73 ± 0.21 | 0.90 ± 0.13 | **<0.001** |
| **Perceived health** | 64.4 ± 20.4 | 81.6 ± 17.2 | **<0.001** |
| **Overall daily activity impairment due to health (except work) (%)** | 52.6 ± 29.9 | 16.4 ± 25.9 | **<0.001** |
| **Mental and social health** |  |  |  |
| **Depression** |  |  | **<0.001** |
| No depression | 456 (43.7%) | 1903 (85.3%) |  |
| Mild depression | 380 (36.4%) | 254 (11.4%) |  |
| Moderate depression | 128 (12.3%) | 56 (2.5%) |  |
| Moderately severe depression | 65 (6.2%) | 12 (0.5%) |  |
| Severe depression | 15 (1.4%) | 6 (0.3%) |  |
| **Coping** | 38.1 ± 9.7 | 40.5 ± 10.6 | **<0.001** |
| **Loneliness** |  |  | **<0.001** |
| Not lonely | 570 (54.6%) | 1524 (68.3%) |  |
| Moderately lonely | 296 (28.4%) | 534 (23.9%) |  |
| Severely lonely | 178 (17.0%) | 173 (7.8%) |  |
| **Lifestyle** |  |  |  |
| **Smoking** |  |  | **<0.001** |
| Never smokers | 505 (48.4%) | 1240 (55.6%) |  |
| Former smoker | 445 (42.6%) | 848 (38.0%) |  |
| Current smoker | 94 (9.0%) | 143 (6.4%) |  |

<sup>a</sup> The sample size for PCC is generally n=1044 and for non-PCC n=2231, unless otherwise noted.
COPD, chronic obstructive pulmonary disease; EQ5D, EuroQol-5 dimension; OI, orthostatic intolerance; PEM, post-exertional malaise; PCC, post-COVID-19 condition; QoL, quality of life; ref., reference group

At the individual level, participants with PCC were more often low- and medium-educated. The PCC group reported poorer health (i.e., more often had PCC at baseline, ≥1 comorbidities, diabetes, cardiovascular diseases, asthma, COPD, fibromyalgia, PEM, OI, mild/severe fatigue, ≥1 grade dyspnea, problems with mobility, self-care, daily activities, pain/discomfort, and anxiety/depression). They also reported a lower quality of life, lower perceived health and greater impairment in performing daily activities due to their health. PCC participants showed more (severe) depression, had lower mean coping score and higher proportion of loneliness. The PCC group included a higher proportion of former and current smokers compared to non-PCC (Table 1).

### 3.2 Comparison of perceived stress between PCC and non-PCC

Adults with PCC had a higher mean perceived stress score (mean ± SE: 20.75 ± 0.27) than adults without PCC (mean ± SE: 15.65 ± 0.19), adjusted for age and gender (p<0.001). Figure 4 shows the adjusted distribution of perceived stress scores, while taking baseline PCC status into account showing that adults who never had PCC had lowest stress levels (mean ± SE: 15.27 ± 0.20), followed by adults recovered from former PCC (mean ± SE: 17.44 ± 0.41), and then followed by both adults with new onset PCC (mean ± SE: 19.83 ± 0.50) and those with persistent PCC (mean ± SE: 21.07 ± 0.30). Using the median PSS-14 score (16) as cut-off, the proportion perceived stress was 64.3% in the PCC group and 39.8% in the non-PCC group.

**Fig. 4.**
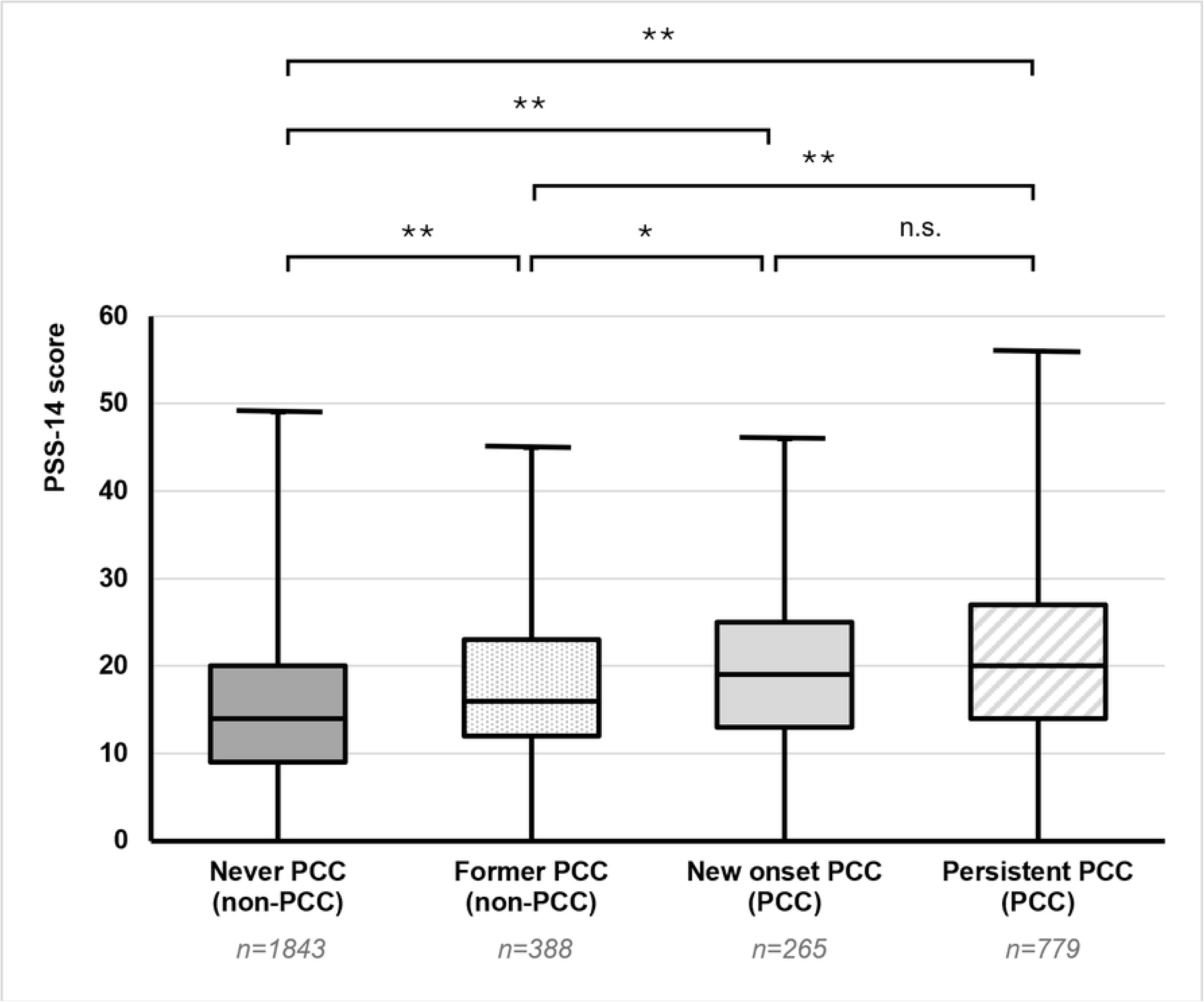
Boxplot of Perceived Stress Scale 14 (PSS-14) score for participants from the PRIME study, who never had post-COVID-19 condition (PCC), former PCC, new onset PCC and persistent PCC. * p=0.001, **p<0.001, n.s. non-significant

### 3.3 Factors associated with perceived stress in PCC and non-PCC (Table 2)

#### 3.3.1 Societal factors

Living in a neighborhood with higher livability and neighborhood cohesion was associated with less stress in PCC.

**Table 2.**
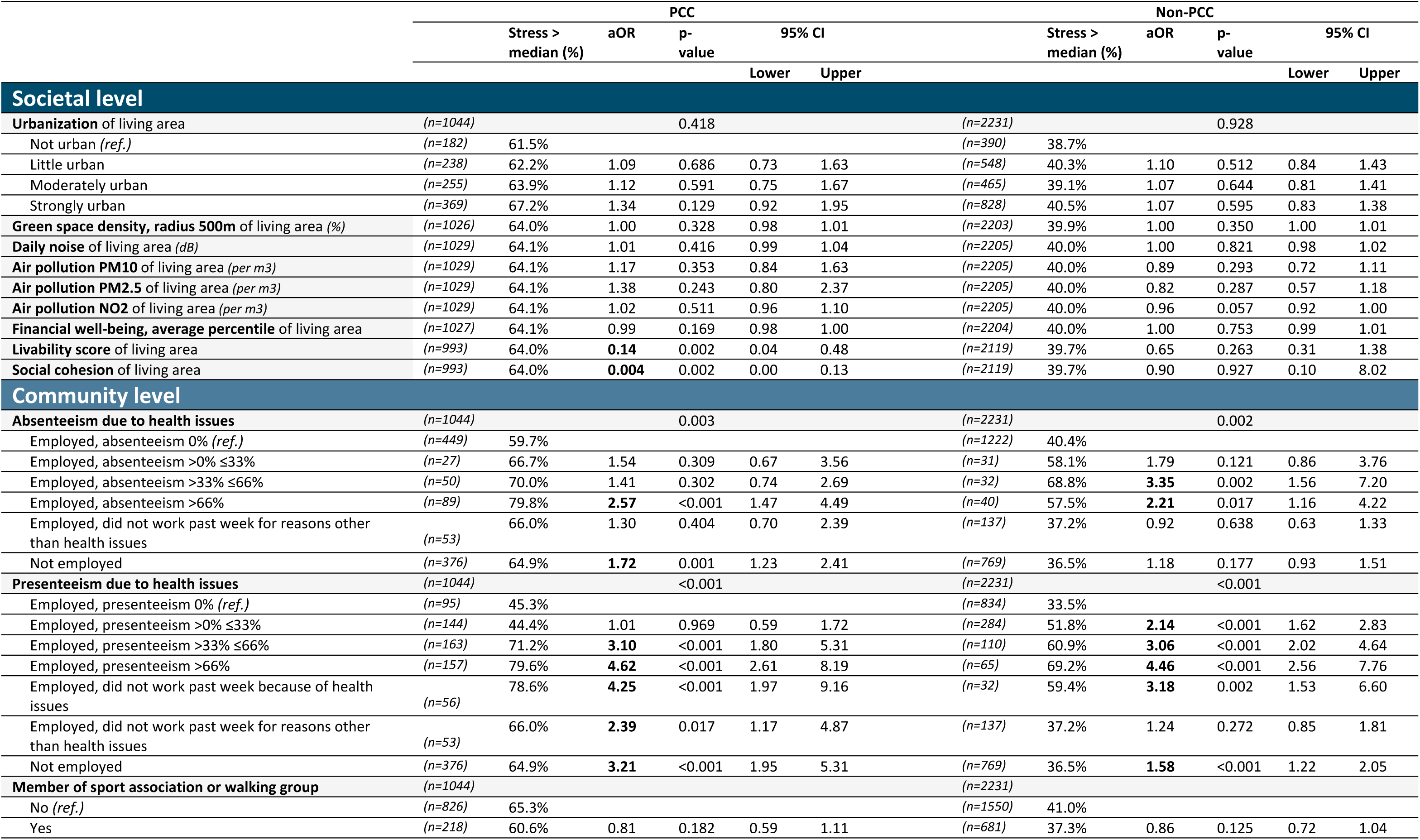
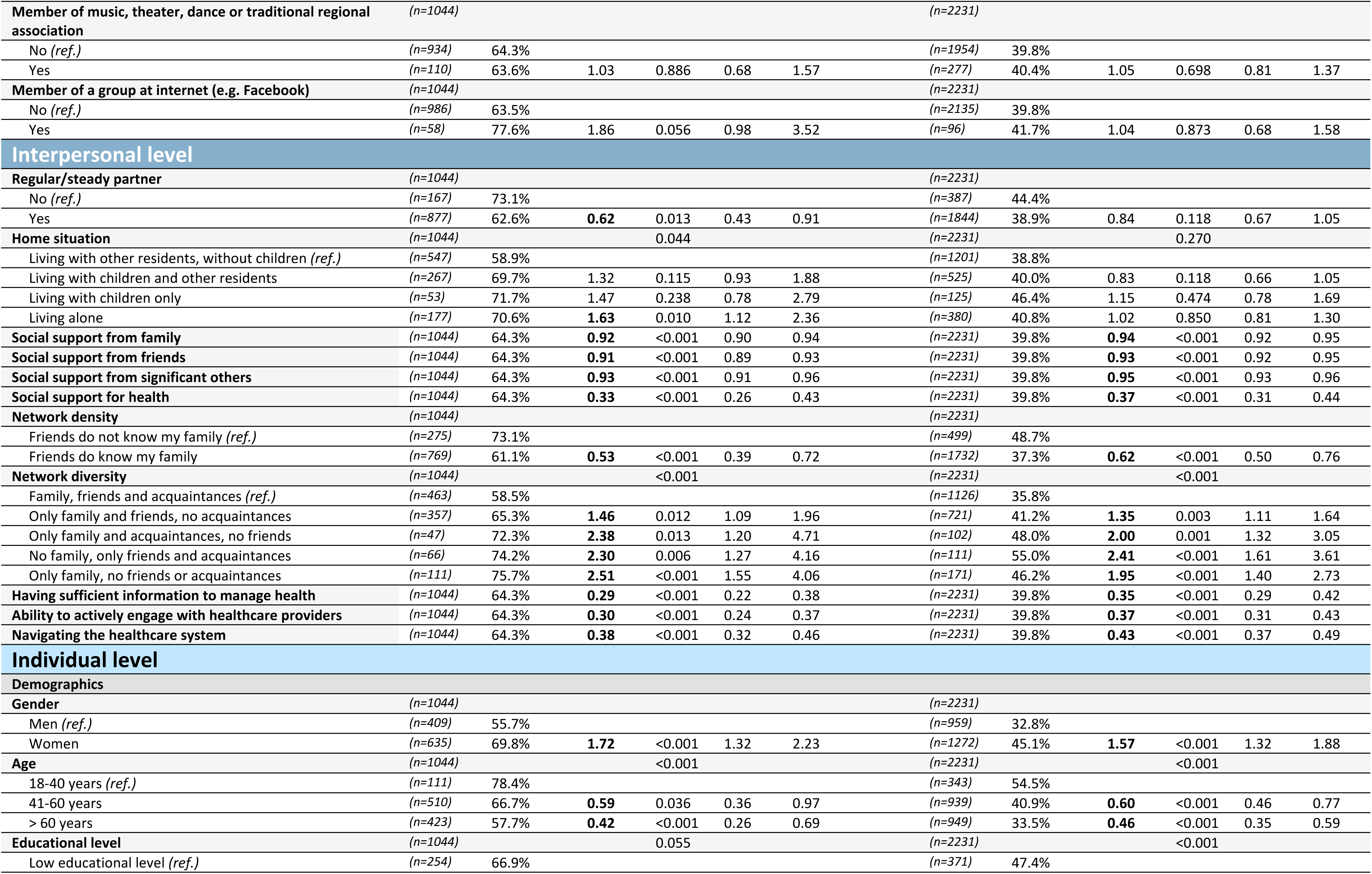
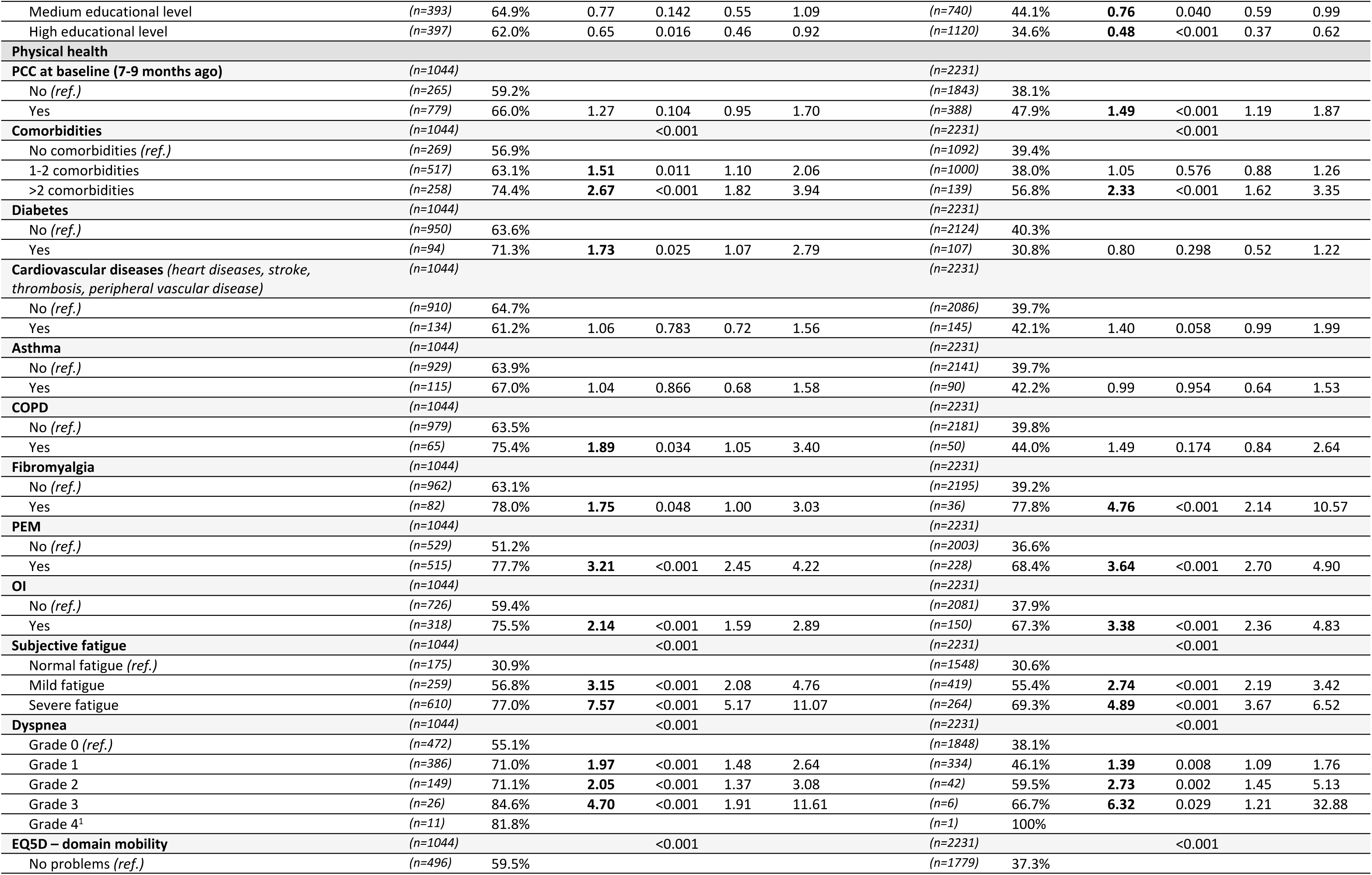
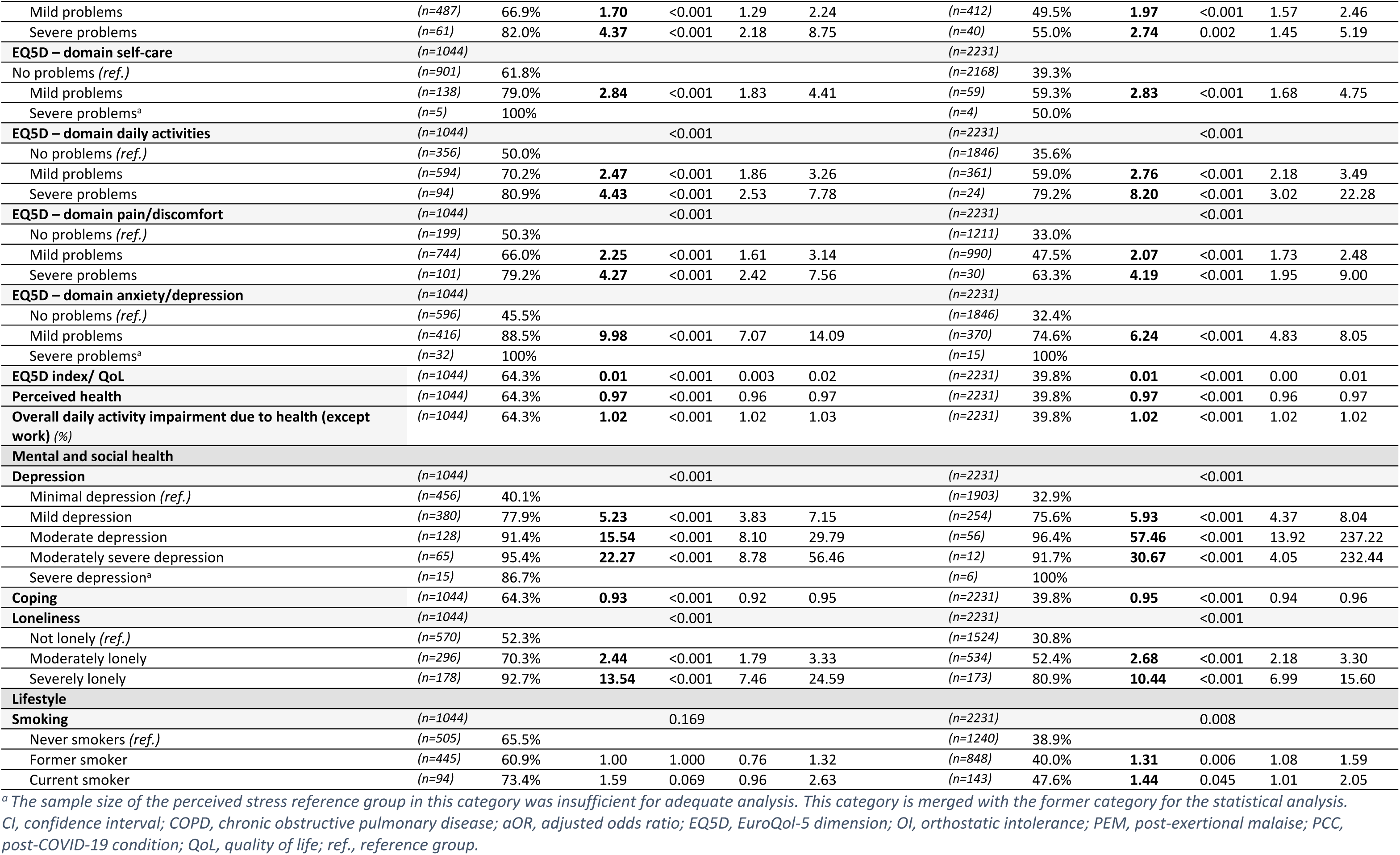
Factors associated with perceived stress above the median (versus ≤ median) in participants from the PRIME study with post-COVID-19 condition (PCC) and without post-COVID-19 condition (non-PCC), using logistic regression adjusted for age and gender.

#### 3.3.2 Community factors

Associated with more stress were absenteeism between >33% and ≤66% (non-PCC group), absenteeism exceeding 66% (both PCC and non-PCC groups), presenteeism up to 33% (non-PCC group), presenteeism above 33% (both groups), not working due to health issues (both groups), and not working due to reasons other than health issues (PCC-group) and unemployment (both groups).

#### 3.3.3 Interpersonal factors

Associated with less stress in both PCC and non-PCC groups were greater social support (from family, friends, significant others), greater health literacy (sufficient information to manage health, receiving social support for health, able to actively engage with healthcare providers and to navigate the healthcare system), and having a more dense and a more diverse network. In the PCC group, having a regular partner was associated with less stress, while living alone was associated with more stress.

#### 3.3.4 Individual factors: demographics and lifestyle

Associated with more stress were being women (both groups), former and current smoker (non-PCC). Associated with less stress were older age (both groups), medium and high education (non-PCC group).

#### 3.3.5 Individual factors: physical health

Associated with more stress in both PCC and non-PCC included having more than two comorbidities, fibromyalgia, PEM, OI, mild and severe subjective fatigue, dyspnea grade 1 and higher, greater impairment in daily activities, as well as having problems with mobility, self-care, daily activities, pain/discomfort and anxiety/depression. Past PCC was also associated with more stress in persons who did not have current PCC (non-PCC group).

Further associated with more stress were having 1-2 comorbidities, diabetes, and COPD (PCC group). Higher QoL and better perceived health were associated with less stress (in both groups).

#### 3.3.6 Individual factors: mental and social health

Depression and loneliness were associated with more stress (both groups), with higher odds of stress for more severe depression and loneliness, whereas coping ability was associated with less stress (both groups).

### 3.4 Heterogeneity of effects

By combining data from the PCC and non-PCC groups and including an interaction term between PCC group (PCC vs. non-PCC) and the stress-associated factors, only a few factors were found to differ between PCC and non-PCC. Factors that showed heterogeneity were livability (p=0.036; associated with stress only in PCC group), social cohesion (p=0.010; only associated in PCC group), presenteeism >0% ≤33% (p=0.014; only associated in non-PCC group), unemployment (p=0.030; stronger associated in PCC group), diabetes (p=0.024; only associated in PCC group), fibromyalgia (p=0.047; stronger associated in non-PCC group), problems with anxiety/depression (EQ5D) (p=0.036; stronger associated in PCC group) and coping ability (p=0.023; stronger associated in PCC group).

### 3.5 Attenuation of perceived stress differences between PCC and non-PCC

The PCC group had a higher odd (adjusted for age and gender) to perceived stress than the non-PCC group (aOR: 2.80, 95% CI: 2.39 – 3.28, p<0.001). When adding the socio-ecological levels, consisting of identified stress-associated factors, one by one to the basic model, the odds ratio progressively attenuated with each level and became non-significant (Table 3 and S3 table).

**Table 3.** Multivariable logistic regression to assess which levels attenuate the higher odds of perceiving stress for the post-COVID condition (PCC) group versus the non-PCC group.

| Model # | Factors included | aOR | p-value | 95% CI |  |
| --- | --- | --- | --- | --- | --- |
|  |  |  |  | Lower | Upper |
| Model 1 | Basic model (age, gender and PCC status) | **2.80** | <0.001 | 2.39 | 3.28 |
| Model 2 | Model 1 + societal factors | **2.75** | <0.001 | 2.34 | 3.22 |
| Model 3 | Model 2 + community factors | **2.04** | <0.001 | 1.72 | 2.43 |
| Model 4 | Model 3 + interpersonal factors | **1.31** | 0.006 | 1.08 | 1.59 |
| Model 5 | Model 4 + individual factors (demographics, lifestyle) | **1.28** | 0.014 | 1.05 | 1.55 |
| Model 6 | Model 5 + individual factors (physical health, part 1) | 0.98 | 0.890 | 0.78 | 1.24 |
| Model 7 | Model 6 + individual factors (physical health, part 2) | 0.84 | 0.144 | 0.66 | 1.06 |
| Model 8 | Model 7 + individual factors (mental and social health) | 0.78 | 0.051 | 0.60 | 1.001 |

361 *aOR, adjusted odds ratio; CI, confidence interval*

## 4.0 Discussion

The results of the current study demonstrated that participants of the PRIME post-COVID study with PCC had higher perceived stress levels (mean ± SE: 20.75 ± 0.27) than those without PCC (mean ± SE: 15.65 ± 0.19). A range of environment (societal, community, interpersonal) and individual factors were associated with perceived stress, including social cohesion and livability of living area, work absenteeism and presenteeism, social support, network diversity, network density, PEM, diabetes and loneliness. Most identified stress-associated factors were similar in PCC and non-PCC, but nearly all these factors were more prevalent in participants with PCC. These findings highlight the multifaceted complexity of stress in PCC, encompassing individual and environmental aspects. Moreover, the higher prevalences of unfavorable characteristics in PCC emphasize the broader impact of PCC.

The current study demonstrated that adults with PCC had higher stress levels than adults without PCC, which is consistent with the majority of previous studies [15–17]. One study (with smaller sample size n=26) showed no difference [43]. The current study further demonstrated that participants with PCC, whether their condition is persistent or new onset, perceived similar levels of stress. Moreover, stress levels decreased after PCC recovery, but remained elevated, in line with a previous study [15].

While in society and clinical practice, perceived stress is regarded as an individual matter, this study highlights the importance of the environment, alongside individual factors.

Identified factors and their effect-sizes were largely similar between the PCC and non-PCC group in our study, indicating that adults with PCC generally do not have a distinct profile of stress-related factors. Rather, participants with PCC more often experienced such factors, which may contribute to the higher stress in PCC.

We will highlight a few factors. At the societal level, participants with PCC more often lived in an environment that is less in line with the needs of the residents (livability) than those without PCC, although there is only a slight difference. We demonstrated, in line with another study, that lower neighborhood social cohesion was associated with stress [44].

Notably, we observed this association, and the link between stress and livability, only in participants with PCC, indicating a mismatch between their needs and neighborhood resources; needs may be higher due to the illness while resources may even be less present or inaccessible.

At the community level, presenteeism (>33%) and extensive absenteeism (>66%) due to health problems were associated with stress, partly in line with previous research demonstrating associations with presenteeism, not absenteeism [45]. The higher prevalence of absenteeism and presenteeism in the PCC group may be due to debilitating PCC symptoms that likely interfere with the ability to maintain regular attendance and productivity.

At the interpersonal level, social network aspects (i.e., lower network diversity, lower density, and less social support) were associated with more stress. Literature suggested that social networks moderate stress [46] and it was striking that participants with PCC in our study had a less favorable social network. The debilitating nature of PCC makes it harder to maintain or build social relationships, and important social network resources may be lost.

At the individual level, various physical, mental and social factors were associated with stress. An example is PEM, which was more prevalent in PCC. PEM might contribute to elevated stress both directly and indirectly through factors such as limited social contacts, reduced ability to work and decreased productivity. Another example is diabetes, which may be accompanied by diabetes distress, defined as ‘the negative emotional or affective experience resulting from the challenge of living with the demands of diabetes’ [18]. The association between diabetes and stress in our study was observed only in participants with PCC, possibly because the added burden of PCC surpassed the participants’ ability to manage diabetes distress. Furthermore, loneliness was associated with stress, aligning with previous research [47], and was more prevalent in the PCC group. This is consistent with previous findings that long-term health conditions increase the risk of loneliness [48].

Altogether, the presence of a diverse range of environmental aspects (i.e. societal, community and interpersonal factors) in addition to individual factors (e.g. physical, mental and social factors) accounted for higher stress in PCC (Table 3). Notably, the community and interpersonal factors appeared to have a substantial impact attenuating the odds ratio (i.e. the difference in stress between the PCC and non-PCC group) by 25% and 36%, respectively, while societal and individual demographical factors attenuated the odds ratio by just 2%.

### 4.1 Implications

The novel insights gained from this study contributed to better understanding of stress in adults with PCC. We recommend that professionals involved in the care of PCC patients, as well as the patients’ social environment, recognize that adults with PCC are more likely to perceive stress, as they are likely to encounter a wide range of stress-associated factors.

Since adults with PCC generally do not have a distinct profile of stress-related factors, the factors targeted in PCC stress treatment may be similar to those for adults without PCC (e.g. improving social networks). However, the approach may need to differ and be tailored to the unique challenges of PCC. For instance, encouraging participation in sports events to build social networks may be suitable for those without PCC but is typically inappropriate or could be harmful for adults with PCC. Strategies aimed to mitigate stress in adults with PCC, need to keep three notes in mind. First, realize that stress involves a complex interplay of a variety of environment and individual factors, possibly also influencing each other, suggesting that a singular focus will be inadequate. For example, in practice, healthcare professionals may focus on improving coping to mitigate stress. However, in our view, adults with PCC encounter stress-associated factors to such an extent that focusing solely on coping is insufficient. Second, acknowledge that even after recovery of PCC, stress is an issue to be addressed. Although PCC may be (partly) resolved, its consequences may not (e.g. reintegrating or finding a job, or rebuilding social network takes time). Third, it is important to acknowledge that identification and subsequent ‘treatment’ of stress may support adults with PCC, as it may offer some relief, but is not to be taken to ‘cure’ PCC.

### 4.2 Strengths and limitations

The major strengths of this study are the large, population-based sample and the design with a comparison group without PCC. The incorporation of the non-PCC group provided crucial insights into whether associated factors were generally associated with stress or specifically in PCC. Second, a wide range of factors across four distinct socio-ecological levels was evaluated, allowing for a thorough assessment of environment to individual aspects involved in perceived stress. Third, the study used perceived stress as outcome measure, which is powerful due to the inclusion of the participants’ subjective appraisal to a potential stressful situation, instead of objectively counting the number of potentially stressful life-events. Finally, the use of validated scales across various measures enhanced reliability and validity of findings.

The main limitation was limited external validity of the findings for severe PCC cases, since the PRIME questionnaires may be too exhaustive for them to complete. We advised completing the questionnaire in multiple sessions to minimize this issue. However, we cannot rule out underrepresentation of individuals with severe PCC, which may have led to underestimation of stress levels in PCC. Additionally, individuals who are less digitally skilled are less represented. Second, we cannot rule out that some adults currently classified as “never PCC” may have had PCC before baseline assessment during the first year and a half of the COVID-19 pandemic. Third, measures were mainly self-reported, thus no biomedical data (e.g. cortisol levels) could be evaluated. If included, these might have provided additional insights into the body’s stress response. Fourth, no conclusions regarding cause and effect can be drawn from the current cross-sectional study, whether stress-associated factors may influence stress levels, be influenced by stress, or act reciprocally with stress, nor can be established whether factors act directly, indirectly, or both. Lastly, while the questionnaire data was collected in 2022, the census tract data corresponded to earlier years. We selected the closest available census data to 2022, though this difference is unlikely to affect the results, since environmental data are relatively stable over time.

### 4.3 Conclusion

To conclude, adults with PCC perceived higher stress levels compared to those without PCC, which may be attributable to the high prevalences of a complex set of environmental (societal, community, interpersonal) and individual stress-associated factors in adults with PCC. The currently obtained knowledge about the occurrence of stress in adults with PCC and understanding of associated aspects contributes to a greater understanding of the multifaceted impacts of PCC.

## 5.0 Statements and Declarations

### 5.1 CRediT author statement

**Senne M.C.E. Wijnen:** Writing – original draft, Methodology, Conceptualization, Formal analysis, Visualization. **Céline J.A. van Bilsen:** Writing – review & editing, Methodology, Conceptualization. **Demi M.E. Pagen:** Writing – review & editing, Conceptualization.

**Annemarie Koster:** Writing – review & editing. **Christian J.P.A. Hoebe:** Writing – review & editing. **Nicole H.T.M. Dukers-Muijrers:** Writing – review & editing, Supervision, Methodology, Funding acquisition, Conceptualization.

### 5.2 Data availability

The datasets presented in this article are not readily available because the data contains potentially identifying patient information. Data are available on request from the head of the data-archiving South Limburg Public Health Service for researchers who meet the criteria for access to confidential data. Requests to access the datasets should be directed to.

## Acknowledgements

We gratefully acknowledge LCJ Steijvers, CDJ den Heijer, M Van Herck, MA Spruit, AW Vaes, CPB Moonen, S Mujakovic, HLG ter Waarbeek, and N Bouwmeester-Vincken for their valuable contribution throughout the shaping and data collection of the PRIME post-COVID study. Furthermore, we gratefully acknowledge all study participants for their valuable contributions, as well as the colleagues and collaborators from AWPG Mosa of the GGD Zuid-Limburg and Maastricht University for their support in facilitating the PRIME study. Geo-data were collected as part of the Geoscience and Health Cohort Consortium (GECCO), which was financially supported by the Netherlands Organization for Scientific Research (NWO), the Netherlands Organization for Health Research and Development (ZonMw), and Amsterdam UMC. More information on GECCO can be found on www.gecco.nl

## 7.0 Supporting information captions

**S1 table.** The 14 items of the Perceived Stress Scale (PSS-14) used in the PRIME study questionnaire.

**S2 table.** Detailed information on census tract data and self-reported data from the PRIME study, as used in the current analysis.

**S3 table.** Multivariable model (model #8 of Table 3) including all factors that were significantly associated with perceived stress in the PCC and/or non-PCC group (n=3112) (PRIME study).

## Notes

### Competing Interest Statement

The authors have declared no competing interest.

### Clinical Trial

N/A, NCT05128695

### Author Declarations

The Medical Ethical Committee of Maastricht University Medical Center+, Maastricht Netherlands waived this study (METC2021-2884), as the Medical Research Involving Human Subjects Act (WMO) did not apply to this study.

